# Forecast of the COVID-19 outbreak and effects of self-restraint in going out in Tokyo, Japan

**DOI:** 10.1101/2020.04.02.20051490

**Authors:** Junko Kurita, Tamie Sugawara, Yasushi Ohkusa

**Affiliations:** Department of Nursing, Tokiwa University, Ibaraki, Japan; National Institute of Infectious Diseases, Tokyo, Japan

**Keywords:** COVID-19, countermeasures, necessary ICU beds, SIR model

## Abstract

**Background:** The number of patients of COVID-19 in Tokyo has been increasing gradually through the end of March, 2020.

**Object:** Support for policymaking requires forecasting of the entire course and outcome of the outbreak if a self-restraint in going out is not initiated. Moreover, the effects of a self-restraint in going out must be considered when choosing to initiate one. Method: Data of Tokyo patients with symptoms during January 14 – March 28, 2020 were used to formulate a susceptible–infected–recovered (SIR) model using three age classes and to estimate the basic reproduction number (R_0_). Based on the estimated R_0_, We inferred outbreak outcomes and medical burden if a self-restraint in going out were not enacted. Then we estimate the self-restraint in going out effects.

**Results:** Results suggest R_0_ as 2.86, with a 95% confidence interval of [2.73, 2.97]. Exhaustion of medical resources can be expected to occur on April 26 if no self-restraint in going out occurs. If a self-restraint in going out were enacted from April 6, and if more than 60% of trips outside the home were restricted voluntarily, then medical care service could be maintained.

**Discussion and Conclusion:** The estimated R_0_ was similar to that found from other studies conducted in China and Japan. Results demonstrate that a self-restraint in going out with reasonable cooperation of residents is required to maintain medical care.

## Introduction

The initial case of COVID-19 in Japan was that of a patient who showed symptoms when returning from Wuhan, China on January 3, 2020. As of March 28, 2020, 1150 cases had been announced as infected in the community of Japan, excluding asymptomatic cases, those for which the onset date or age was not reported, those infected abroad, and those infected on a large cruise ship: the Diamond Princess [1].

In metropolitan Tokyo, which has approximately 14 million residents, 234 symptomatic cases were identified as of March 28, 2020. The entire course of the outbreak must be predicted to evaluate the necessary medical resources for policymaking. Moreover, one must evaluate, as a worst case scenario, exhaustion of medical resources which can occur when medical needs far exceed the capacity of medical resources. Especially, the capacity of intensive care unit (ICU) facilities is usually not so large. They are expected to be allocated quickly to patients.

To forecast these phenomena, we construct a simple susceptible–infected–recovered (SIR) model for Tokyo incorporating the necessary medical resources. Then we predict whether exhaustion of medical resources occurs.

## Method

We applied a simple SIR model [2,3,4] with three age classes: children 19 years old or younger, adults 20–59 years old, and elderly people 60 years old or older. We assumed some protection of children [5]: 40% ([20, 70] %) of children were protected infection [4]. The incubation period was assumed to be equal for all people of the three age classes and following the empirical distribution inferred for the outbreak in Japan.

Experiences of Japanese people living in Wuhan until the outbreak provide information related to mild cases because complete laboratory surveillance was conducted for them. During January 29 – February 17, 2020, 829 Japanese people returned to Japan from Wuhan. Each had undergone a test to detect COVID-19; of them, 14 were found to be positive for COVID-19 [6]. Of those 14, 10 Japanese people had exhibited mild symptoms; the other 4 showed no symptom. Moreover, two Japanese residents of Wuhan exhibited severe symptoms: one was confirmed as having contracted COVID-19. The other died, although no fatal case was confirmed as COVID-19 by testing. In addition, two Japanese residents of Wuhan with mild symptoms were refused re-entry to Japan even though they had not been confirmed as infected. If one assumes that the Japanese fatal case in Wuhan and that the two rejected re-entrants were infected with COVID-19, then 2 severe cases, 12 mild cases, and 4 asymptomatic cases were found to exist among these Japanese residents of Wuhan. We therefore apply these proportions of asymptomatic cases to symptomatic cases in the simulation.

We also assumed that the degrees of infectivity among the severe patients and mild patients were equal and asymptomatic cases has half of infectiousness in the symptomatic cases were assumed. This assumption about relative infectiousness among asymptomatic cases compared with symptomatic cases was used also in simulation studies for influenza [7–11].

We sought to ascertain R_0_ to fit the number of patients during 14 January – 28 March and to minimize the sum of squared residuals among the reported numbers and the fitted values. Its 95% confidence interval (CI) was calculated using 10,000 iterations of bootstrapping for the empirical distribution of epidemic curves.

Contact patterns among children, adults, and elderly people were estimated in an earlier study [12]. We identified the following contact patterns: the share of children contacting with other children accounted for 15/19 of all contacts, contact with adults accounted for 3/19, and contact with elderly people accounted for 1/19; the share of adults contacting children accounted for 3/9 of all contacts; those contacting with other adults were 5/9, and those contacting with elderly people were 1/9; elderly people contacting with children were 1/7, with adults were 2/7, and with other elderly people were 4/7.

We assumed that contact frequencies in the same age class decreased in the same proportion in all age classes if the Tokyo Metropolitan Government were to declare a self-restraint in going out in Tokyo. However, contact frequencies among other age classes were assumed not to be changed by a self-restraint in going out because most of the contact with other age classes can be presumed to occur at home. Contacts at home will probably be unaffected by a self-restraint in going out.

Experience in Japan has revealed the pneumonia incidence in elderly COVID-19 patients as 28%. That among adults is 20% based on national data in Japan until March 23 [1]. We used these ratios to assume ratios of severe cases to symptomatic cases. Among children, no pneumonia case has been reported. Only pneumonia cases were received hospital treatment. The length of hospitalization was assumed as 30 days. Of those pneumonia cases, 30% were assumed to require the use of an intensive care unit (ICU) for 20 days.

However, if patients requiring care at an ICU cannot receive it, then we assumed that the CFR among them was 100%. We define the exhaustion of medical resources as circumstances under which the necessary ICU bed number becomes greater than 70% of all existing ICU beds. In Tokyo, there are 1000 ICU beds. Therefore, if the necessary number of ICU beds becomes greater than 700, medical service at the facilities will face crisis.

We used data of the COVID-19 community outbreak of patients in Japan who showed any symptom during January 14 – March 28, 2020 in Tokyo. We excluded some patients who had been infected abroad and who returned from abroad and those who were presumed to be infected persons from the Diamond Princess. They were presumed not to be community-acquired in Japan.

Published information about COVID-19 patients with symptoms from the Ministry of Labour, Health and Welfare (MLHW) Japan or TMG was usually affected adversely by some delay because of uncertainty during onset to visiting a doctor or in the timing of a physician’s suspicion of COVID-19. Therefore, published data of patients must be adjusted at least a few days. To adjust the data, we applied the following regression analysis. We set *Xt-k|t* as the number of patients for whom the onset date was *t-k* published on day *t*. The dependent variables are the degree of reporting delay, *Xt-k-m|t* / *Xt-k-m|t-m*, where *k*>*m* for several *m* and *k*. Here, *m* denotes the difference of the publishing dates between the data published on different date. Date *t* represents the publishing date of the latest publishing. The explanatory variables were 1/*k*, 1/*m*, and 1/*km*. The degree of reporting delay was estimated as [estimated coefficient of constant term] + [estimated coefficient of 1/*k*]/*k*, when *m* was sufficiently large and time had passed. Therefore, this estimated degree of reporting delay multiplied by the latest published data is expected to be a prediction of the number of patients for whom the onset date was *t*-*k*. We used this adjusted number of patients in the latest few days. We used published data of 2, 5, 6 and 9–17 March, 2020 provided by MLHW [1].

First, we estimated R_0_. Then we predicted the peak date, total number of symptomatic cases, maximum number of newly infected symptomatic cases per day, beds, and ICU beds. We also predicted the date of exhaustion of medical resources. Moreover, we predicted the effects of the self-restraint in going out from April 6 and measured whether the medical system would be maintain. We calculated the 95% CI through 10,000 bootstrapped distributions.

## Ethical consideration

All information used for this study has been collected under the Law of Infection Control, Japan. There is therefore no ethical issue related to this study.

## Results

During January 14–March 28 in Tokyo, 4 cases among children, 145 cases among adults, and 85 cases among elderly people were identified as community-acquired COVID-19 for whom the onset date was published. Figure 1 depicts the empirical distribution of incubation period among 62 cases for which the exposed date and onset date were published by MHLW. Its mode and median were six days; the average was 6.74 days.

**Figure 1:**
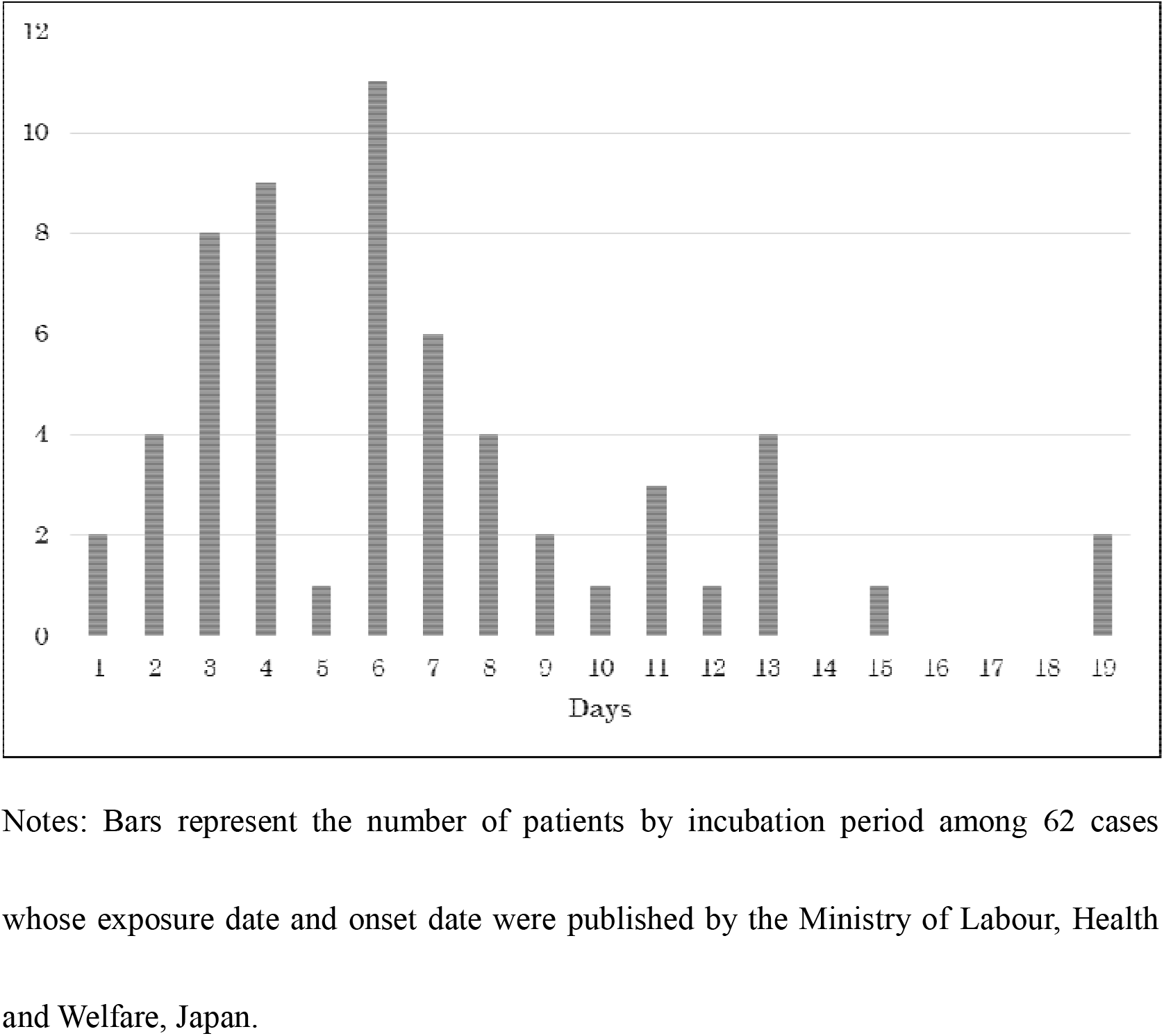
Empirical distribution of incubation period published by Ministry of Labour, Health and Welfare, Japan (number of patients)

Figure 2 depicts epidemic curves published for 2, 6, 10, 12 and 14 March. From this information, we estimated the degree of reporting delay. Those results are presented in Table 1. The table shows that 1*/k*, 1/*m*, and 1/*km* are all significant. When *m* is sufficiently large, the effects of 1/*m* and 1/*km* converge to zero. Therefore, the estimated degree of reporting delay consists of the term of 1/*k* and a constant term. Based on these results, we predict the degrees of reporting delay as 19.3 for *k*=1, 9.64 for *k*=2, 6.42 for *k*=3, and so on.

**Figure 2:**
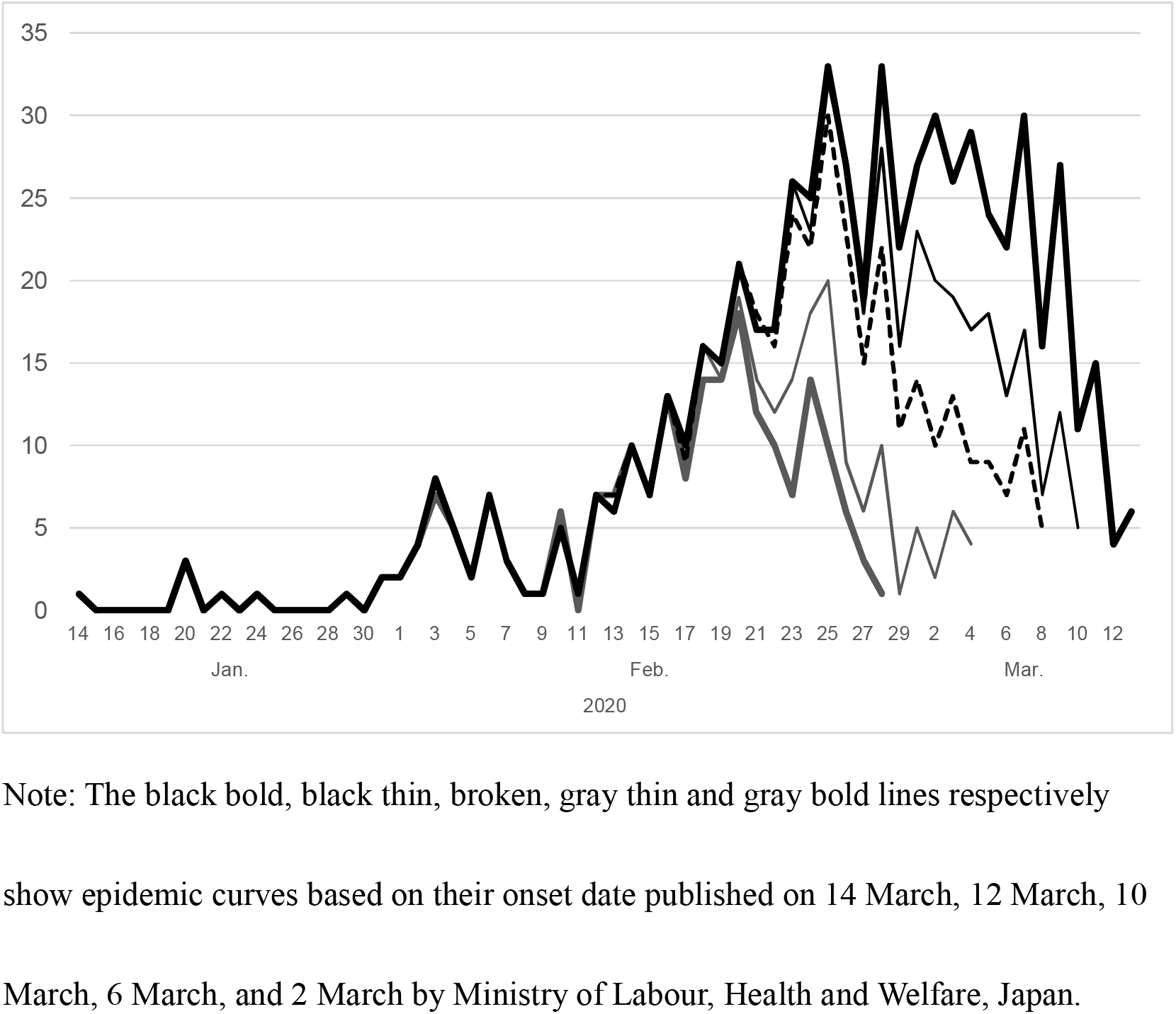
Epidemic curves of COVID-19 in Japan published on 2, 6, 10, 12 and 14 March, 2020 by Ministry of Labour, Health and Welfare, Japan (number of patients)

**Table 1:** Estimation results of the degree of reporting delay

| | Estimated coefficient | $p$-value | 95%CI | |
| --- | --- | --- | --- | --- |
|  |  |  | Lower bound | Upper bound |
| $1/k$ | 16.9 | .000 | 14.7 | 19.0 |
| $1/m$ | 1.01 | .025 | 0.140 | 2.05 |
| $1/(km)$ | -21.8 | .000 | -26.7 | -16.9 |
| Constant | 0.266 | .236 | -0.175 | 0.707 |

Note: The number of observations was 323. The coefficient of determination was 0.457.
$k$ denotes the number of days until the last reported day. $m$ denotes the difference between the publishing date in the past and the most recent publishing date. The dependent variable is the among of reporting delay, which is defined as $X_{t-k-m}/t / X_{t-k-m}/t-m$ , where $X_{t-k}/t$ is the number of patients for whom the onset date was $t-k$ published on day $t$ . We used the number of patients by onset day published on 2, 5, 6, 10, 11, 12, 13, and 14 March 2020 by the Ministry of Labour, Health and Welfare, Japan.

The R_0_ values were estimated as 2.86 and 95%CI [2.73, 2.97]. Outcomes, presented as Table 2, show that the total number of patients with any symptom was estimated as 6.41 [6.38, 6.47] million. The maximum number of patients with new onset was estimated as 0.117 [0.115, 0.120] million per day at the peak. At the peak, the maximum number of the administered patients with pneumonia was estimated as 0.776 [0.766, 0.796] million per day. Moreover, the maximum number of patients who need an ICU bed was estimated as 0.180 [0.180, 0.187] million per day. Finally, results show that 70% of ICU capacity will be exceeded on 28 [26, 29] April. Results of the self-restraint in going out are presented in Table 3 according to the proportion of self-restriction of people leaving their home. The exhaustion of medical resources could be prevented if more than 60% of trips outside of the home were restricted. The exhaustion of medical resources could postponed by three months if more than 50% but fewer than 60% of trips outside of the home going were restricted.

**Table 2:** Prediction of patients and necessary medical resources in Tokyo (in million)

|  | Median | 95% confidence interval (CI) |  |
| --- | --- | --- | --- |
|  |  | Lower | Upper |
| Total number of symptomatic patients | 6.41 | 6.38 | 6.47 |
| Maximum number of newly symptomatic patients per day | 0.117 | 0.115 | 0.120 |
| Maximum number of inpatients | 0.776 | 0.766 | 0.796 |
| Maximum number of necessary intensive care unit (ICU) patients | 0.180 | 0.180 | 0.187 |
| Date of exhaustion of medical resources | April 28 | April 26 | April 29 |

Note: This table presents simulation results that can be expected without self-restraint in going out. 95% CI was estimated both through 10,000 iterations of bootstrapping. “exhaustion of medical resources” was defined as necessary ICU beds greater than 70% of capacity.

**Table 3:** Predicted effects of self-restraint in going out in Tokyo as measured by date of exhaustion of medical resources

| Proportion of self-restraint<br>in going out (%) | Estimation | 95% confidence interval (CI) |  |
| --- | --- | --- | --- |
|  |  | Lower | Upper |
| 0 | 28 April | 26 April | 29 April |
| 5 | 30 April | 28 April | 1 May |
| 10 | 2 May | 20 April | 3 May |
| 15 | 5 May | 2 May | 6 May |
| 20 | 8 May | 5 May | 10 May |
| 25 | 12 May | 9 May | 14 May |
| 30 | 17 May | 13 May | 20 May |
| 35 | 25 May | 20 May | 28 May |
| 40 | 5 June | 29 May | 8 June |
| 45 | 24 June | 14 June | 29 June |
| 50 | 30 July | 13 July | 8 August |
| 55 | 20 October | 3 September | 21 November |
| >60 | No exhaustion of medical resources |  |  |

Note: “exhaustion of medical resources” is defined as necessary intensive care unit beds greater than 70% of capacity.

## Discussion

We applied a simple SIR model with three age classes including asymptomatic cases and assuming some proportion of children as protected. An earlier study [13–15] estimated R_0_ for COVID-19 as 2.24–3.58 in Wuhan. Our obtained R_0_ of 2.86 was very similar. However, one study revealed that R_0_ in Japan up through 26 February was just 0.6 [16]. It means that large outbreak will not emerge in Japan except for sporadic cases and medical resources will have never exhausted. It was undoubtedly not correct even at the time of their investigation and the responsibility to insist on contact tracing to detect cluster was not light.

Moreover, on April 26, medical services can be expected to be critical. Results also demonstrate that if a self-restraint in going out is initiated on April 6 and if more than 60% of trips outside the home were restricted voluntarily, then medical care could be maintained. It is noteworthy that no law exists to enforce curfews in Japan. Therefore, a self-restraint in going out ask, not force, residents to avoid leaving their home voluntarily. Consequently, cooperation with a self-restraint in going out must achieve voluntary cooperation to a great degree. Evidence related to compliance is scarce because no lockdown has been conducted in Japan to date. An exceptional study asked people about restricting movement outside the home if the government asked them to do it [17]. Results showed that 93.3% would comply voluntarily with such a government request. The exhaustion of medical resources due to COVID-19 might be avoided in Japan, even with no enforcement of a lockdown, because strong compliance can be expected.

Our unpublished research suggests that school closure since March 2 decreased contact frequencies among children by 40%. Voluntary event cancellation since February 27 decreased it among adults by 50%. Although a exhaustion of medical resources has been postponed, it appears to be unavoidable if we apply these numbers to the effect of self-restraint in going out.

The SIR model is too simple to incorporate households, firms or schools. It is a completely mixed model. It therefore ignores some difference inside and outside of those groups. It can adjust contact patterns to mimic some policies including self-restraint in going out or school closures as in the present study. A model highlighting differences inside and outside of those groups is an individual-based model (IBM), which mimics movements and contacts of individuals. It can therefore evaluate behavioral changes of individuals directly [6–9,17]. Therefore, we must use IBM for evaluation of a self-restraint in going out instead of a SIR model. No IBM exists for COVID-19, but an IBM exists for pandemic influenza. Especially, the most precise IBM, RIBM, has been developed in Japan using actual data of transportation [17, 18]. It indicated that 60% voluntary restriction to going out can reduce prevalence by 40 percentage points for pandemic flu [18]. Therefore, it shared the same result that 60% voluntary restriction out of trips outside the home can avoid exhaustion of medical resources with the present study for COVID-19.

## Conclusion

We predicted outbreak of COVID-19 and effects of self-restraint in going out in Tokyo. We estimated a exhaustion of medical resources in late April in Tokyo if a self-restraint in going out were not applied. If a self-restraint in going out were enacted from April 6, and if more than 60% of trips outside the home were restricted voluntarily, then medical care service could be maintained. Such a self-restraint in going out might avoid a exhaustion of medical resources. However, it is noteworthy that such a self-restraint in going out might continue until a vaccine for COVID-19 can be developed. Costs of a self-restraint in going out would be huge if it were to last for more than a half of year. Its cost-effectiveness is expected to represent a concern in this case. This study represents the authors’ opinion. It does not reflect any stance of our affiliation.

## Data Availability

Japan Ministry of Health, Labour and Welfare. Press Releases of Domestic Situation (in Japanese)

https://www.mhlw.go.jp/stf/seisakunitsuite/bunya/0000121431_00086.html

## Acknowledgments

We acknowledge the great efforts of all staff at public health centers, medical institutions, and other facilities who are fighting the spread and destruction associated with COVID-19.

